# Effects of Six Weeks of Supervised Telerehabilitation on Pulmonary Function, Functional Capacity, and Dyspnoea among individuals with Long COVID

**DOI:** 10.1101/2023.09.27.23296254

**Authors:** Alya Al Mheiri, Srilatha Girish, Sampath Kumar Amaravadi

**Author notes:** **Corresponding author:** Srilatha Girish, Associate Professor, Department of Community Physiotherapy, MGM Institute of Physiotherapy, Aurangabad, Maharashtra, India. Department of Community Physiotherapy, MGM Institute of Physiotherapy, Aurangabad, Maharashtra, India. Department Physiotherapy, Institute of Sport, Nursing and Allied Health, University of Chichester, West Sussex, PO196PE. United Kingdom.

## Abstract

**Background:** Long COVID, characterized by persistent symptoms post-COVID-19, presents challenges like reduced functional capacity, pulmonary function, and dyspnea. Telerehabilitation, a remote healthcare approach, is gaining attention for its potential to address these issues

**Objectives:** The study aims to determine the effect of telerehabilitation on functional capacity by six-minute walk test (MWT), pulmonary function by pulmonary function test (PFT), dyspnea by modified medical research council (mMRC) and level of physical activity by Global physical activity questionnaire (GPAQ) in individuals with long COVID.

**Materials and Methods:** At Al Ain Hospital, UAE, a 6-week telerehabilitation program for Long COVID patients aged 18-75 was studied using single-group pretest-posttest quasi-experimental design. Pre and post assessments included 6MWT, PFT, mMRC, and GPAQ. The study also monitored technical issues and session adherence.

**Results:** Participants had an average age of 49.30 ± 15.46, height of 163.80 ± 9.76, and weight of 78.70 ± 15.58, with a gender ratio of 12 females to 8 males. After six weeks of telerehabilitation, significant improvements were seen in the 6MWT (21% improvement) and PFT (4% FVC increase, 8% FEV1 increase, 1% FEV1/FVC increase, and 11% PEF increase). mMRC scale scores post-rehabilitation were significantly lower, indicating substantial improvement in dyspnea levels with clinical significance.

**Conclusion:** This approach has shown tangible benefits in enhancing functional capacity, pulmonary function, reducing dyspnea, and improving physical activity levels among individuals with Long COVID. The results of the study demonstrate the feasibility and effectiveness of implementing a telerehabilitation program for individuals with Long

## BACKGROUND

Coronavirus 2019 (COVID-19), caused by the outbreak of the acute respiratory syndrome coronavirus 2 (SARS-CoV-2) is a global pandemic and has led to an unprecedented public health crisis. [1] According to the World Health Organization (WHO), 1,038,228 cases of COVID-19 have been observed in United Arab Emirates (UAE). [2] The clinical spectrum of COVID-19 varies from asymptomatic infection to severe outcomes including respiratory illness, multiorgan failure, and fatal cases. [3,4] Clinical manifestations have been categorized into three phases by the joint guidelines proposed by the National Institute for Health and Care Excellence (NICE), the Scottish Intercollegiate Guidelines Network (SIGN), and the Royal College of General Practitioners (RCGP): ‘Acute COVID-19’ (signs and symptoms of COVID-19 infection up to 4 weeks), ‘ongoing symptomatic COVID-19’ (from 4 weeks up to 12 weeks), and ‘post-COVID-19 syndrome’ (when signs and symptoms continue beyond 12 weeks).[5]

The majority (∼ 80%) symptomatic individuals recover without hospitalization. Approximately15% become seriously ill and require oxygen, while the remaining 5% become critically ill and need intensive care. [6] While most individuals recover from COVID-19, survivors have reported persistent symptoms that remain unresolved for months after being discharged from the hospital. [7] Various terms, such as “long haulers,” “Chronic COVID syndrome,” “post-COVID-19 syndrome,” “post-acute COVID-19,” or “Long-COVID,” have been used interchangeably to describe the lingering effects of COVID-19. Among these, “Long COVID” is widely accepted globally. [8]

While a specific definition is yet to be universally agreed upon, Long COVID is generally described as a cluster of new symptoms that emerge around 12 weeks after a confirmed or suspected case of COVID-19, in the absence of no other clinical diagnosis. [9] Individuals experiencing Long COVID commonly report impaired lung functions, post-exertional fatigue, and dyspnea. Additionally, other symptoms include neurocognitive impairment, autonomic dysfunction, and mental health issues like anxiety and depression, among others. [10]

Long COVID can manifest regardless of the severity of the initial infection, hospitalization status, or age. [11] Long COVID can lead to decreased physical activity levels and have a significant impact on the overall quality of life. [12,13].

Research has shown that dedicated efforts to curb the spread of COVID-19, including the implementation of prevention methods, development of treatments, and distribution of vaccines, have been instrumental in reducing the initial stage of infections. [14] Shifting the perspective on the pandemic and giving significant attention to Long COVID is crucial. Various professional organizations have emphasized the importance of early detection and rehabilitation for individuals experiencing Long COVID symptoms. [10] Despite ongoing research, the mechanisms behind Long COVID remain incompletely understood, and there is currently limited evidence for effective pharmacological management. As a result, exercise training has emerged as a viable alternative for effectively controlling the symptoms associated with Long COVID.

A large body of research has established exercise training as the cornerstone for addressing dyspnea and fatigue in various respiratory conditions in hospital setup (in or patient rehabilitation [15,16] However, training in a hospital setup is challenged by factors including but not limited to low participant uptake, insufficient attendance by the individual, and high dropout rates, accessing hospital-based training are transportation issues, acute exacerbation of the conditions, disruption of daily routine, and symptom severity. As an alternative approach the American Thoracic Society (ATS)/European Respiratory Society (ERS) now recommends telerehabilitation (TR) to extend the benefits of hospital set up. [16,17]

TR is a method of providing rehabilitation services to individuals remotely, utilizing information and communication technologies. [18] Unlike hospital-based rehabilitation, exercising at home may enhance effectiveness, facilitate long-term integration of exercise routines into daily life, and improve compliance. The beneficial impact of exercise training through TR on exertional tolerance and dyspnea in conditions such as COPD has been extensively documented.[18] Nevertheless, there is limited literature available regarding the impact of telerehabilitation training on exertional tolerance and dyspnea in individuals with Long COVID. [19-26] Apart from the challenges faced in hospital-based training, the ongoing pandemic necessitates clinical and public measures to minimize the risk of viral transmission. TR programs present a practical alternative in such situations. The objective of the study was to determine the effect of six weeks of telerehabilitation on pulmonary function, functional capacity and dyspnea in individuals with Long COVID.

## METHOD

### Study design

The current study adopted one group pretest-posttest quasi-experimental design.

### Selection criteria

Participants of both the gender aged between 18 to 75 years discharged from the Physical Medicine and Rehabilitation Department in Al Ain hospital, with a history of molecular testing confirmed COVID-19 infection, having a score of 0-2 on the modified medical research council (mMRC) and having the access and ability to use virtual conference call application (google meet or micro-soft team) were considered for the current study. The participant with mMRC dyspnea score of 3–4, resting heart rate over 100 bpm, uncontrolled hypertension (resting BP > 160/100mmHg), uncontrolled diabetes (e.g., diabetes with random blood glucose >16.7 mmol/L, hemoglobin A1C >7.0%), cerebrovascular disease within six months, intra-articular drug injection or surgical treatment of lower extremities within six months and those who are unable to walk independently without the assistive device were excluded from the study as they require close supervision and not considered fit for telerehabilitation program.

### Sample size calculation

The minimal important difference (MID) for COVID-19 has not been established, a MID of 30 meters is recommended for a 6-minute walk test in chronic diseases. Given the limited knowledge about the long-term course of COVID-19, a MID of 50 was assumed. With 80% power, 5 % alpha error, and an attrition rate of 20%, the required sample was 26.

### Outcome measure

- Lung Functions: Lung functions were measured from a pulmonary function test (PFT) using a portable pulmonary function device (EasyOne Pro® LAB, USA) according to guidelines of the American Thoracic Society. Changes in baseline parameters of vital capacity (FVC), forced expiratory volume in the first second (FEV1), the ratio of FEV1 and FVC (FEV1/FVC), and Peak Exploratory Flow (PEF) at six weeks were assessed. [27]
- Functional capacity: The functional capacity was evaluated by a six-minute walk test (6MWT) in accordance with guidelines from the American Thoracic Society. The change in distance walked in the 30-meter corridor from baseline functional capacity at six weeks was assessed. [28-29]
- Dyspnea perception: The individual’s perception of dyspnea was measured with the Modified Medical Research Council Dyspnea Scale (mMRC). Change in baseline perception of dyspnea level at six weeks was assessed on a 5-point ordinal scale (0: no dyspnea; 4: Severe dyspnea). [30]
- Level of Physical Activity: The level of physical activity was measured using the Global physical activity questionnaire (GPAQ) at six weeks of exercise training. [31]

### Procedure

The study was conducted in the physical medicine and rehabilitation department at Al Ain hospital. Ethical approval was obtained Institutional Review Board (Ref. no. (IRB/COHS/STD/04/Jan-2022) of Gulf Medical University and SEHA, Al Ain hospital (Ref no. ERC-19022-1-2022). Consent to recruit the participants from the present study was obtained from Al Ain hospital. To locate the potential participants, the discharge registry of the Physical Medicine and Rehabilitation Department in Al Ain hospital was accessed. Participants were contacted over the phone and the study protocol was explained. Upon obtaining verbal consent, participants were provided with appointments in the hospital for further evaluation of eligibility criteria. On meeting the eligibility criteria, participants signed informed consent form.

The virtual platform used in the current study was google meet. Google meet is a web-based video conferencing application from Google. The user interface is extremely simple, easy to use, and intuitive. The researcher ensured that all the participants met the minimum requirements for google meet (meet mobile application, Gmail mobile application, or a supported web browser, a compatible device with minimum system requirements, a supported operating system, broadband connection to the internet, a built-in web camera or external USB camera) video conference. All the participants were trained for using the google meet interface in terms of starting and joining video meetings; adjust meeting’s audio and video setup.

Baseline assessment of all the selected participants were carried out for functional capacity, pulmonary function and dyspnea using 6MWT, PFT and mMRC scale respectively. Following this all the participants received two sessions of face-to-face training on setting up the virtual platform, individualized exercise training to be carried out at participants residence. In addition, participants were trained to monitor heart rate and SPO2 through a portable oximeter, and rate of perceived exertion (RPE) by modified Borg scale throughout the intervention. A schedule of home-based exercise training was provided for all the participants based on their convenience.

A home-based aerobic training was carried out for six weeks with three synchronous sessions per week remotely monitored by the physiotherapist through the google meet platform. Each session lasted for 40 minutes. Each session has a warm-up (10 minutes), conditioning (20 minutes), and cool-down phase (10 minutes). At the beginning of the session, the individual wears a portable oximeter and records HR and SPO2.

The warm-up and cool-down period included breathing exercises as well as stretching exercises of the upper and lower extremities (pectorals, triceps, biceps, deltoid, hamstrings, quadriceps, gastrocnemius muscles) with 30 secs hold of 3 repetitions with intensity to the point of the feeling of slight discomfort).

The conditioning phase consisted of aerobic training. The aerobic training was performed in the form of spot marching (2 minutes) with alternate chest trunk mobilization (1 minute) and dynamic exercises in large muscle groups 2-3 sets of 10 repetitions. The exercise intensity was based on the HRR determined by Karvonen’s formula and was scheduled to increase from 30-40% HRR at the beginning of the session to 40-60% HRR in the final two weeks.

Participants were advised to perform physical activity (swimming, brisk walking, jogging) for 30 minutes on the reminder days. And they were provided with a daily log sheet to monitor compliance with to exercise program. Following six weeks of exercises, appointments were scheduled, and a post-intervention assessment of the outcome measure was taken for functional capacity, pulmonary function and dyspnea.

## RESULTS

A total of 50 participants were contacted by phone after being identified through the Physical Medicine and Rehabilitation Department’s discharge register at Al Ain Hospital. Twenty-four of the 27 participants who consented to participate in current study, presented for further evaluation of eligibility requirements. All of the selection criteria were met by 20 participants and same completed six weeks aerobic training through telerehabilitation. (Figure 1) The demographic characteristics of the participants are shown in Table 1. The mean age of all the participants was 49.30 ± 15.46 years. Average height and weight of the participants in the study were 163.80 ± 9.76 cm and 78.70 ± 15.58 kg, respectively. The study participants were predominantly female (n=12) and nine were with comorbidity of obesity

**Table 1:**
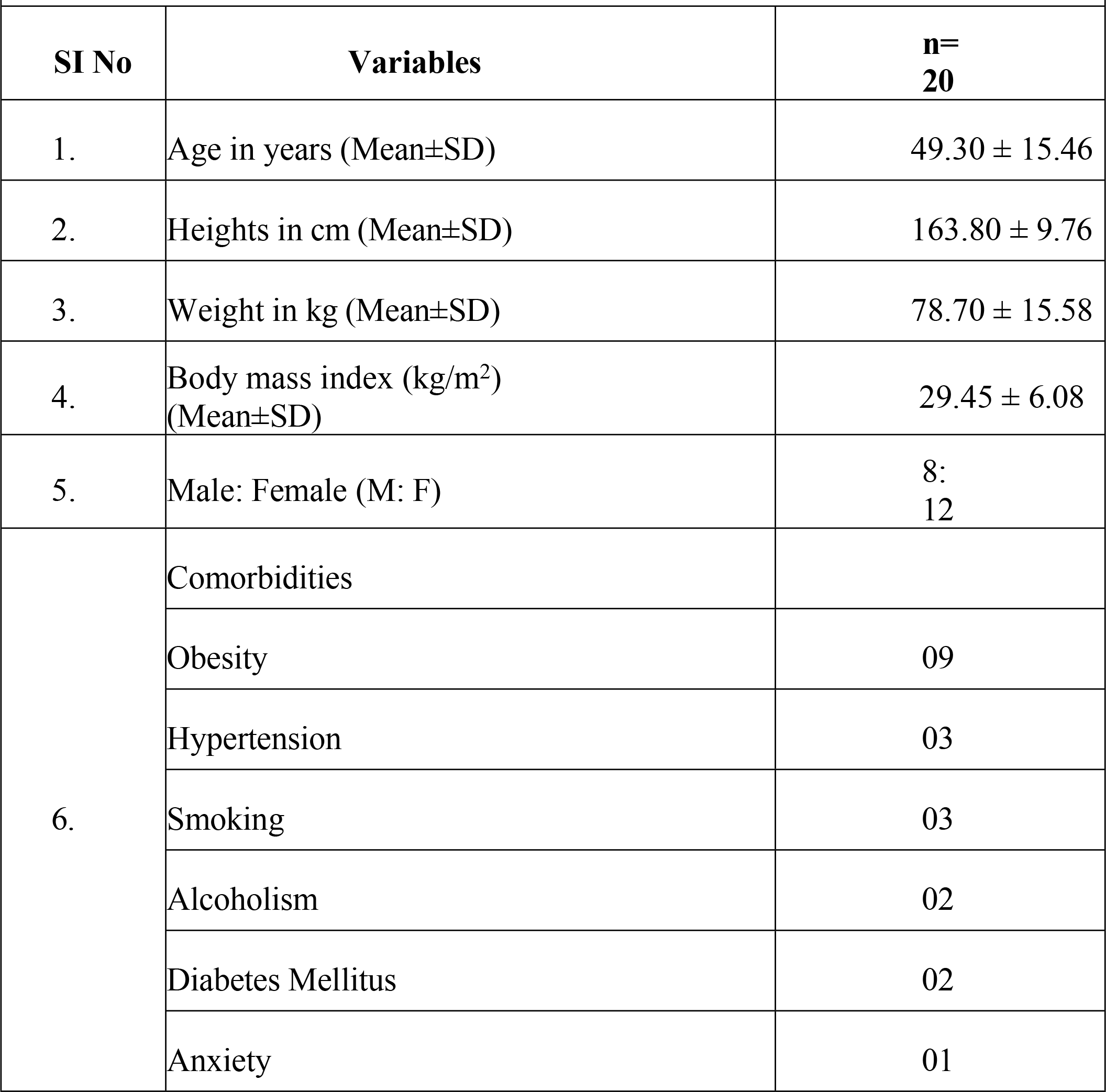
Demographic and clinical charaxcteristics of the study participants.

Table 2 presents the outcomes of the 6MWT, PFT, and mMRC scale scores before and after the telerehabilitation intervention. Overall, the telerehabilitation program demonstrated a statistically significant improvement in the 6MWT and PFT results after a 6-week duration. The 6MWT showed a noteworthy improvement of 21%, while the PFT values exhibited improvements in forced vital capacity (FVC) by 4%, forced expiratory volume in one second (FEV1) by 8%, FEV1/FVC ratio by 1%, and peak expiratory flow (PEF) by 11 units. Moreover, the post-rehabilitation scores of mMRC indicated a significant decrease when compared to the pre-rehabilitation scores.

**Table 2:** Pre and post tele-rehabilitation changes of six-minute walk distance, pulmonary function & dyspnea levels of the study participants.

| Outcome measures |  | Mean | Mean % change | Std. Deviation | Std. Error Mean | Correlation | P value Sig. (<0.05) |
| --- | --- | --- | --- | --- | --- | --- | --- |
| **Six-MWD (meters)** | Pre | 441.00 | 21.08 | 89.78 | 20.07 | 0.88 | < 0.001 |
|  | Post | 534.00 |  | 95.16 | 21.28 |  |  |
| **PFT (%)** | Pre_FVC | 3.15 | 4.44 | 0.86 | 0.19 | 0.99 | < 0.001 |
|  | Post_FVC | 3.29 |  | 0.84 | 0.18 |  |  |
|  | Pre_FEV1 | 2.50 | 8 | 0.91 | 0.20 | 0.84 | < 0.001 |
|  | Post_FEV1 | 2.70 |  | 0.67 | 0.15 |  |  |
|  | Pre_FEV1/FVC | 83.05 | 1.05 | 5.74 | 1.28 | 0.95 | < 0.001 |
|  | Post_FEV1/FVC | 82.17 |  | 4.75 | 1.06 |  |  |
|  | Pre_PEF | 6.31 | 11.5 | 2.13 | 0.47 | 0.97 | < 0.001 |
|  | Post_PEF | 7.04 |  | 1.93 | 0.43 |  |  |
| **mMRC** | Pre | 2.30 | -47.8 | 0.47 | 0.10 | 0.76 | < 0.001 |
|  | Post | 1.20 |  | 0.41 | 0.09 |  |  |

Note: MWD: Minute walk test(MWD), Forced vital capacity (FVC), forced expiratory volume in the first second (FEV1), Peak Exploratory Flow (PEF) and Modified Medical Research Council (mMRC)

Table 3 provides a comprehensive overview of the statistical significance of changes in the 6MWT, PFT and mMRC scale of the study participants. These results help evaluate the effectiveness of the intervention and provide insights into the impact on functional capacity, pulmonary function and dyspnea in participant The physical activity levels of the participants at baseline were determined using the Global Physical Activity Questionnaire. The findings revealed that 35% (n=7) of the participants were identified as physically active (> 600 MET-min/week), while 65% (n=13) were classified as physically inactive (<600 MET-min/week) at baseline

**Table 3:** Paired T test of six-minute walk distance, pulmonary function & dyspnea levels of the study participants.

| Outcome measures |  | Paired Differences |  |  |  |  |  |  |  |
| --- | --- | --- | --- | --- | --- | --- | --- | --- | --- |
|  |  | Mean | Std. Deviation | Std. Error Mean | 95% Confidence Interval of the Difference | 95% Confidence Interval of the Difference | t | df | Sig. (2-tailed) |
|  |  |  |  |  | Lower | Upper |  |  |  |
| **Six-MWD** | Pre - Post | -93.00000 | 45.54928 | 10.18513 | -114.31772 | -71.68228 | -9.131 | 19 | .000 |
| **PFT** | Pre_FVC - Post_FVC | -.13800 | .11701 | .02616 | -.19276 | -.08324 | -5.275 | 19 | .000 |
|  | Pre_FEV1 - Post_FEV1 | -.19600 | .49637 | .11099 | -.42831 | .03631 | -1.766 | 19 | .093 |
|  | Pre_FEV1/FVC - Post_FEV1/FVC | .88000 | 1.86677 | .41742 | .00632 | 1.75368 | 2.108 | 19 | .049 |
|  | Pre_PEF - Post_PEF | -.72400 | .49180 | .10997 | -.95417 | -.49383 | -6.584 | 19 | .000 |
| **mMRC scale** | Pre -Post | 1.10000 | .30779 | .06882 | .95595 | 1.24405 | 15.983 | 19 | .000 |

Note: MWD: Minute walk test(MWD), PFT: Pulmonary Function Test, Forced vital capacity (FVC), forced expiratory volume in the first second (FEV1), Peak Exploratory Flow (PEF) and Modified Medical Research Council (mMRC)

## DISCUSSION

Telerehabilitation has gained significant popularity and widely promoted during COVID-19 pandemic to assure rehabilitation continuity. However, there is a paucity of published reports in UAE to support the usefulness of telerehabilitation among Long COVID individuals. The current study is the first study in the UAE, conducted with rigorous methodology and ensure safety among UAE individuals with Long COVID.[32]

The findings from current study demonstrated that the telerehabilitation is effective in significantly improving functional capacity, pulmonary function and reducing dyspnea among long COVID individuals. The literature evidenced, exercise delivered via telerehabilitation has led to increase in functional capacity as assessed by the 6 MWT [22–24,26] with low level of certainty. However, its effects on dyspnea [22–24] and pulmonary functions [22,26] were inconclusive.

The current study demonstrated that aerobic training through remote monitoring appears to be same with no incidence of adverse events. One trial has reported adverse events such as chest tightness, weakness, dizziness, sputum discharge, dizziness, chest pain and back pain.[22]

During the pandemic telerehabilitation has been promoted across the globe for health care delivery for various conditions. However, there is a lack of strong scientific evidence on effectiveness of aerobic training on main symptoms of Long COVID specifically with no evidence in UAE. Existing trails presented heterogeneity in terms of participant’s demography, stage of COVID-19 and method adopted in telerehabilitation [22–24,26]

Furthermore, telerehabilitation offers the advantage of delivering therapy wherever is most convenient for the individual, while allowing therapists to prescribe various interventions and receive feedback through modern digital technology. Additionally, individuals can participate in telerehabilitation sessions from the comfort of their own homes or care centers, reducing the burden and involvement of healthcare providers compared to traditional face-to-face setups.

The study demonstrates notable strengths. Firstly, during the implementation of exercises through telerehabilitation, no adverse events were reported, indicating a high level of safety. Secondly, the study observed an impressive adherence rate of over 90% among participants, indicating a strong commitment to attending the exercise sessions. These findings underscore the study’s robustness and highlight the safe implementation and high acceptance of telerehabilitation for delivering exercise interventions.

The current study has few drawbacks. Due to time constraints, participants recruitment was confined to a single center. Furthermore, the lack control group, small sample size, overrepresentation of female and lack of participant’s knowledge on web and technology literacy were also few factors. Hence while generalizing the results of the current study, one must exercise caution. Additionally, these considerations also make determining the intervention’s effectiveness in compared to spontaneous recovery difficult.

The clinical implications of the study are promising. According to preliminary findings from the study, telerehabilitation appears to be a beneficial and convenient method in assisting the recovery of individuals with long COVID. Increasing the number and duration of supervised sessions may reduce the risk of complications associated with long COVID thereby facilitating the improvement in the overall quality of life.

Future research endeavors should place emphasis on implementing a comprehensive monitoring system that accurately tracks the dosage, adherence, and precision of each exercise component. Additionally, it is crucial to consider the long-term effect of the telerehabilitation on Long COVID, an aspect that was not addressed in the present study.

## CONCLUSION

A six-week of aerobic exercise training delivered through telerehabilitation can effectively improve functional capacity, pulmonary function and reduce dyspnea among long COVID individuals. Telerehabilitation, with its high adherence rate and little side effects, makes aerobic delivery possible and well tolerated by individuals with Long COVID. Large scale research is warranted to explore the long-term benefits and optimal implementation strategies of telerehabilitation among Long COVID individuals.

## Data Availability

All relevant data are within the manuscript and its Supporting Information files.

